# High-throughput Genetic Clustering of Type 2 Diabetes Loci Reveals Heterogeneous Mechanistic Pathways of Metabolic Disease

**DOI:** 10.1101/2022.07.11.22277436

**Authors:** Hyunkyung Kim, Kenneth E. Westerman, Kirk Smith, Joshua Chiou, Joanne B. Cole, Timothy Majarian, Marcin von Grotthuss, Josep M. Mercader, Soo Heon Kwak, Jaegil Kim, Jose C. Florez, Kyle Gaulton, Alisa K. Manning, Miriam S. Udler

**Affiliations:** Diabetes Unit, Massachusetts General Hospital, Boston, MA 02114, USA; Broad Institute of MIT and Harvard, Cambridge, MA 02142, USA; Center for Genomic Medicine, Massachusetts General Hospital, Boston, MA 02114, USA; Clinical and Translational Epidemiology Unit, Massachusetts General Hospital, Boston, MA 02114, USA; Department of Pediatrics, University of California San Diego, San Diego, CA 92161, USA; Department of Medicine, Harvard Medical School, Boston, MA 02115, USA; Division of Endocrinology and Center for Basic and Translational Obesity Research, Boston Children’s Hospital, Boston, MA 02115, USA; Takeda Pharmaceuticals, Cambridge, MA 02139, USA; Department of Internal Medicine, Seoul National University Hospital, Seoul 03080, Republic of Korea; GlaxoSmithKline, Cambridge, MA 02140, USA

## Abstract

**Aims/hypothesis:** Type 2 diabetes (T2D) is highly polygenic and influenced by multiple biological pathways. Rapid expansion in the number of T2D loci can be leveraged to identify such pathways, thus facilitating improved disease management.

**Methods:** We developed a high-throughput pipeline to enable clustering of T2D loci based on variant-trait associations. Our pipeline extracted summary statistics from genome-wide association studies (GWAS) for T2D and related traits to generate a matrix of 324 variant x 64 trait associations and applied Bayesian Non-negative Factorization (bNMF) to identify genetic components of T2D. We generated cluster-specific polygenic scores and performed regression analysis in an independent cohort (N=25,419) to assess for clinical relevance.

**Results:** We identified ten clusters, replicating the five from our prior analysis as well as novel clusters related to beta-cell dysfunction, pronounced insulin secretion, and levels of alkaline phosphatase, lipoprotein-A, and sex hormone-binding globulin. Four clusters related to mechanisms of insulin deficiency, five to insulin resistance, and one had an unclear mechanism. The clusters displayed tissue-specific epigenomic enrichment, notably with the two beta-cell clusters differentially enriched in functional and stressed pancreatic beta-cell states. Additionally, cluster-specific polygenic scores were differentially associated with patient clinical characteristics and outcomes. The pipeline was applied to coronary artery disease and chronic kidney disease, identifying multiple shared genetic pathways with T2D.

**Conclusions/interpretation:** Our approach stratifies T2D loci into physiologically meaningful genetic clusters associated with distinct tissues and clinical outcomes. The pipeline allows for efficient updating as additional GWAS become available and can be readily applied to other conditions, facilitating clinical translation of GWAS findings. Software to perform this clustering pipeline is freely available.

## Introduction

Type 2 Diabetes (T2D) has variable contributions of insulin resistance and beta cell dysfunction, and influenced by multiple risk factors, including genetics [1]. Untangling the heterogeneity of T2D may be fundamental to improving patient management and facilitating precision medicine.

Hundreds of loci associated with T2D have been identified in large-scale genetic studies, including genome-wide association studies (GWAS), however, the translation of these established T2D genetic loci into improved understanding of disease pathophysiology has been challenging, due in large part to the variants being non-protein coding [2–5]. Recent studies have leveraged a growing number of available GWAS datasets to connect genetic loci to mechanistic pathways by clustering loci based on shared patterns of associations across multiple traits [2, 6–8]. In our previous work [6], clustering was performed on 94 T2D variants identified by manual curation of published T2D GWAS manuscripts. Soft clustering analysis with Bayesian Non-negative Matrix Factorization (bNMF) of the associations of these 94 T2D variants with 47 diabetes-related traits identified five distinct clusters, recognizable as biological pathways of T2D. A similar set of five clusters of T2D loci were independently identified by Mahajan *et al*., along with a sixth cluster of “undetermined” physiological impact[2]. Of these five shared clusters, two related to beta-cell dysfunction, and the other three clusters represented different mechanisms of insulin resistance: obesity-mediated, abnormal lipodystrophy-like fat distribution, and altered hepatic lipid metabolism [9].

With new T2D loci continuously being discovered and additional GWAS trait summary statistics becoming publicly available, we sought to expand our prior work which involved manual curation of a smaller set of T2D loci. We developed a high-throughput pipeline to enable extraction of hundreds of genetic variants and traits from multiple GWAS to be used for cluster analysis in order to identify new genetic pathways of disease.

## Material and methods

### Pipeline for input variant-trait association matrix for clustering

An overview of preprocessing steps for variants and traits used for generating the input matrix for variant-trait association clustering analysis is illustrated in **Figure S1** with additional details in the **Supplementary Methods**. To obtain a comprehensive set of independent genetic variants associated with T2D, we extracted 21,666 variants reaching genome-wide significance (*P* < 5×10^−8^) from multiple large-scale T2D studies [2, 3, 10–14] in the AMP-Common Metabolic Disease Knowledge Portal (CMDKP) [15] (**Table S1**) and performed stringent LD-pruning of variants at r^2^ < 0.1 (**Table S2**) as well as filtering and removal of multi-allelic, ambiguous (A/T or C/G), or poorly represented (<80%) of trait GWAS datasets.

For trait selection, we utilized summary statistics available for 75 GWAS of glycemic traits, anthropometric traits, vital signs, and additional laboratory measures in the AMP-CMDKP or UK Biobank [15, 16] (**Table S3, Supplementary Methods**). Our goal was to let the genetics guide which traits were included in the clustering analysis, and thus traits were used only if the minimum *P*-value across the final set of variants was lower than a Bonferroni *P*-value cutoff of 0.05/N_final_variants (N=324). We then removed highly correlated traits (with |r| ≥ 0.85) to reduce redundancy.

For the selected lists of variants and traits, we utilized the GWAS summary statistics to generate a matrix of standardized Z-scores, choosing the T2D risk-increasing allele for each variant and dividing the estimated regression coefficient beta by the standard error. To account for the differences in sample size across trait GWAS studies, we scaled the standardized Z-scores by dividing by the square root of the sample size for each variant in each trait GWAS and then multiplied by the mean of square root of median sample size across all SNPs in each GWAS.

This pipeline was also used for coronary artery disease (CAD) and chronic kidney disease (CKD) with six CAD GWAS [17–19] and 39 CKD-related GWAS [11, 16, 20–23] queried.

### bNMF clustering

The variant-trait association matrix Z (m by n, m: # of variants, n: # of traits) was constructed as above. We then generated a non-negative input matrix X (2m by n) by concatenating two separate modifications of the original Z matrix: one containing all positive standardized Z-scores (zero otherwise) and the other all negative standardized Z-scores multiplied by -1. The bNMF procedure factorizes X into two matrices, W (2m by K) and H^T^ (n by K), as X ∼ WH with an optimal rank K, corresponding to the association matrix of variants and traits to the number of clusters, respectively (**Supplementary Methods**) [6]. The key features for each cluster are determined by the most strongly associated traits, a natural output of the bNMF approach. bNMF algorithm was performed in R Studio for 1,000 iterations with maximum number of cluster K set to 20. To define a set of strongest-weighted variants in each cluster, we employed a method to determine a cluster weight cutoff that maximized the signal to noise ratio of weights (**Figure S2**).

### Cluster associations with relevant phenotypes using GWAS summary statistics

We generated GWAS-partitioned polygenic scores (GWAS pPS) for each cluster utilizing inverse-variance weighted fixed effects meta-analysis of GWAS summary statistics including the set of strongest-weighted variants above the weight cutoff for each cluster using the dmetar package in R [24] (**Supplementary Methods**). For testing T2D cluster associations with cardiometabolic outcomes, the significance threshold was set to 0.05/(7×K), representing a Bonferroni correction for K clusters and 7 outcomes (**Table S4**).

### Functional annotation and enrichment analysis

At each locus, we calculated approximate Bayes Factors (aBF) for all variants 500 kb upstream and downstream with r^2^ ≥ 0.1, with the index variant (100% credible set) from effect size estimates and standard errors, using the approach of Wakefield [25]. We then calculated a posterior probability for each variant by dividing the aBF by the sum of all aBF in the credible set. We obtained previously published 13-state ChromHMM [26] chromatin state calls for 28 cell types, excluding cancer cell lines [27]. We also compiled candidate cis-regulatory elements (cCREs) for 14 cell types and subtypes from published single cell chromatin accessibility datasets [28, 29]. We assessed enrichment of annotations within clusters by overlapping 100% credible set variants for signals in each cluster with cell type epigenomic annotations (chromatin states and cCREs). We also assessed epigenomic enrichment in single cell pancreatic tissue using a second method. As previously described [30], we subset loci from the Beta-cell 1 and 2 clusters, annotated variants using cCREs from INS^high^ and INS^low^ beta cells, and applied fgwas [31] in the fine mapping mode (**Supplementary Methods**).

### Partitioned Polygenic Score (pPS) analysis in the Mass General Brigham Biobank

The Mass General Brigham (MGB) Biobank (formerly Partners Biobank) includes more than 120,000 consented patients across the MGB healthcare system [32, 33] (**Supplementary Methods**). Approval for data analysis was obtained by the MGB IRB, study 2016P001018. We employed algorithmically defined outcome phenotypes for T2D, CAD, and CKD developed by the MGB Biobank [34]. Additional phenotypic data (laboratory measures, vital signs, and anthropometric measures) were extracted from outpatient clinic encounters between 2015-2020, and median values were generated. Up to 36,000 samples were genotyped using three versions of the Biobank SNP array offered by Illumina, which underwent harmonization and rigorous quality control (**Supplementary Methods**). Phasing was performed with SHAPEIT [35] and then imputed with the Haplotype Consortium Reference Panel [36] using the Michigan Imputation Server [37]. All SNPs were genotyped or imputed with high quality (r^2^ values > 0.95).

We performed individual-level analyses on individuals of European ancestry based on self-reported ancestry and genetic PC’s, totaling 25,419 individuals. T2D partitioned polygenic scores (pPSs) for each cluster were generated by multiplying a variant’s genotype dosage by its cluster weight, with only the top-weighted variants included, as defined above. Logistic and linear regression were performed in R v3.6.2, adjusting for age, sex, and PC’s.

## Results

### Ten T2D genetic clusters identified by high-throughput approach

We employed a novel high-throughput pipeline to enable extraction of loci from GWAS summary statistics files and generate a variant-trait association input matrix for clustering analysis (**Figure S1**). Our pipeline started with summary statistics from 13 T2D GWAS studies available in the AMP-CMDKP [15], from which we extracted 21,666 variants associated with T2D reaching genome-wide significance and performed stringent LD-pruning and optimization to identify 324 T2D variants, representing independent T2D risk loci (see **Materials and methods**). We let the T2D genetics guide selection of 64 relevant traits to include in subsequent cluster analysis, such that each trait was significantly associated with at least one T2D variant. Soft clustering of the resulting 324 by 64 variant-trait association matrix was performed using bNMF.

The plurality of bNMF iteration results converged on ten clusters (36.3%), which captured the five clusters identified in our previous work [6] as well as five novel clusters (**Table S5, S6**). The remaining bNMF iterations converged on nested clusters, with 6 clusters in 0.3%, 7 clusters in 1.1%, 8 clusters in 8.3%, 9 clusters in 26.6%, 11 clusters in 22.6%, 12 clusters in 4.4% and 13 clusters in 0.4%. The same six clusters (Beta-cell 1, Beta-cell 2, Proinsulin, Obesity, Lipodystrophy, Liver/Lipid, as described below) were identified in all iterations, based on inspection of constituent variants and traits.

To interpret these clusters, we examined their most highly weighted loci and traits, as well as the aggregate associations of cluster loci with the traits via GWAS pPS (**Table 1, Table S7, Figure S3**). The clusters were named after their most defining traits. Four of the clusters (Beta-cell 1, Beta-cell 2, Proinsulin, and Lipoprotein A) appeared to represent mechanisms of insulin deficiency, with T2D risk-increasing alleles in each cluster collectively associated with reduced fasting insulin and reduced homeostatic assessment of beta cell function (HOMA-B; GWAS pPS *P*-values<0.05). Another five clusters (Obesity, Lipodystrophy, Liver/Lipid, ALP negative, Hyper Insulin Secretion) reflected mechanisms of impaired insulin action, with the T2D risk alleles in these clusters associated with increased fasting insulin and homeostatic assessment of insulin resistance (HOMA-IR; GWAS pPS *P-*values<0.05). The remaining cluster (SHBG) was driven by one T2D allele which was not significantly associated with fasting insulin, but trended toward increased levels (**Table 1, Figure 1, Figure S3, Table S7**).

**Table 1.** Overview of T2D clusters. Physiological impact, key traits, key loci, suspected mechanism for each cluster. The numbers in parentheses next to cluster names indicate the numbers of top-weighted variants in each of the clusters.

| Cluster | Physiological impact | Key traits | Key loci | Suspected mechanism | Note |
| --- | --- | --- | --- | --- | --- |
| Beta-cell 1 (63) | Insulin deficiency | CIR (-), DI (-) | *MTNR1B, CDKAL1, HHEX, C2CD4A, ANK1, ST6GAL1, SLC35C1, SLC30A8* | Beta cell function, glucose homeostasis | Beta Cell cluster from Udler et al. 2018 divided into 2 clusters |
| Beta-cell 2 (28) |  | fasting proinsulin adj FI (+), HOMA-B (-), fasting insulin (-) | *TCF7L2, SLC30A8, ADCY5, GCK, DGKB, MTNR1B, C2CD4A* | Beta cell function, insulin processing | Beta Cell cluster from Udler et al. 2018 divided into 2 clusters |
| Proinsulin (18) |  | fasting proinsulin adj FI (-), HOMA-B (-) | *ARAP1, STARD10* | Insulin synthesis | replicated Proinsulin cluster from Udler et al. 2018 |
| Lipoprotein A (1) |  | lipoprotein A (+) | *SLC22A3/LPA* | Lipoprotein A metabolism | new cluster in this study |
| Obesity (35) | Insulin resistance | BMI (+), waistC (+), % body fat (+), CRP (+) | *FTO, MC4R* | Obesity-mediated insulin resistance | replicated Obesity cluster from Udler et al. 2018 |
| Lipodystrophy (54) |  | adiponectin (-), ISI (-), HDL (-) | *IRS, PPARG, KLF14* | Fat distribution-mediated insulin resistance | replicated Lipodystrophy cluster from Udler et al. 2018 |
| Liver/Lipid (11) |  | CRP (-), TG (-), GGT (-) | *GCKR, HNF1A, PPP1R3B, TOMM40, PNPLA3* | Liver/Lipid metabolism | replicated Liver/Lipid cluster from Udler et al. 2018 |
| ALP negative (4) |  | ALP (-) | *ABO* | Alkaline phosphatase activity levels | new cluster in this study |
| Hyper Insulin Secretion (32) |  | DI (+), CIR (+) | *PPP1R3B, CNTN2, DTNB, TNF, SREBF1* | Insulin secretion, inflammation | new cluster in this study |
| SHBG (1) |  | SHBG (-) | *SHBG* | SHBG metabolism | new cluster in this study |

**Figure 1.**
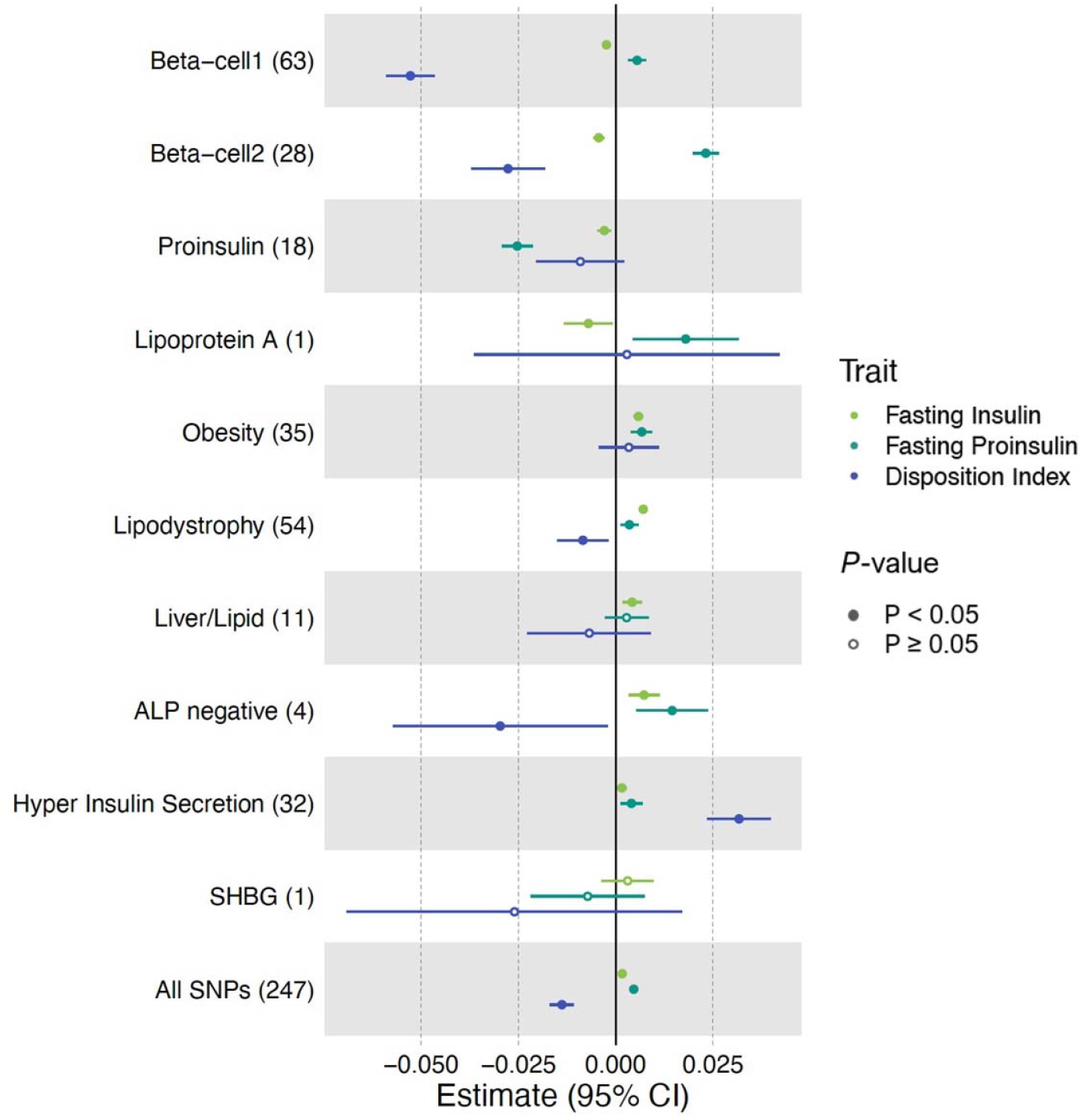
Cluster associations with metabolic traits using GWAS. Standardized effect sizes with 95% confidence intervals of cluster pPS-trait associations derived from GWAS summary statistics shown in forest plot. Three metabolic traits (Fasting Insulin, Fasting proinsulin adjusted for fasting insulin, Disposition Index) that help discriminate clusters are displayed. The numbers in the parenthesis next to cluster names indicate the number of variants included in the analysis in each cluster. “All SNPs” include all the variants that are top-weighted in at least one cluster. Filled points indicate *P*-values less than 0.05.

**Figure 2.**
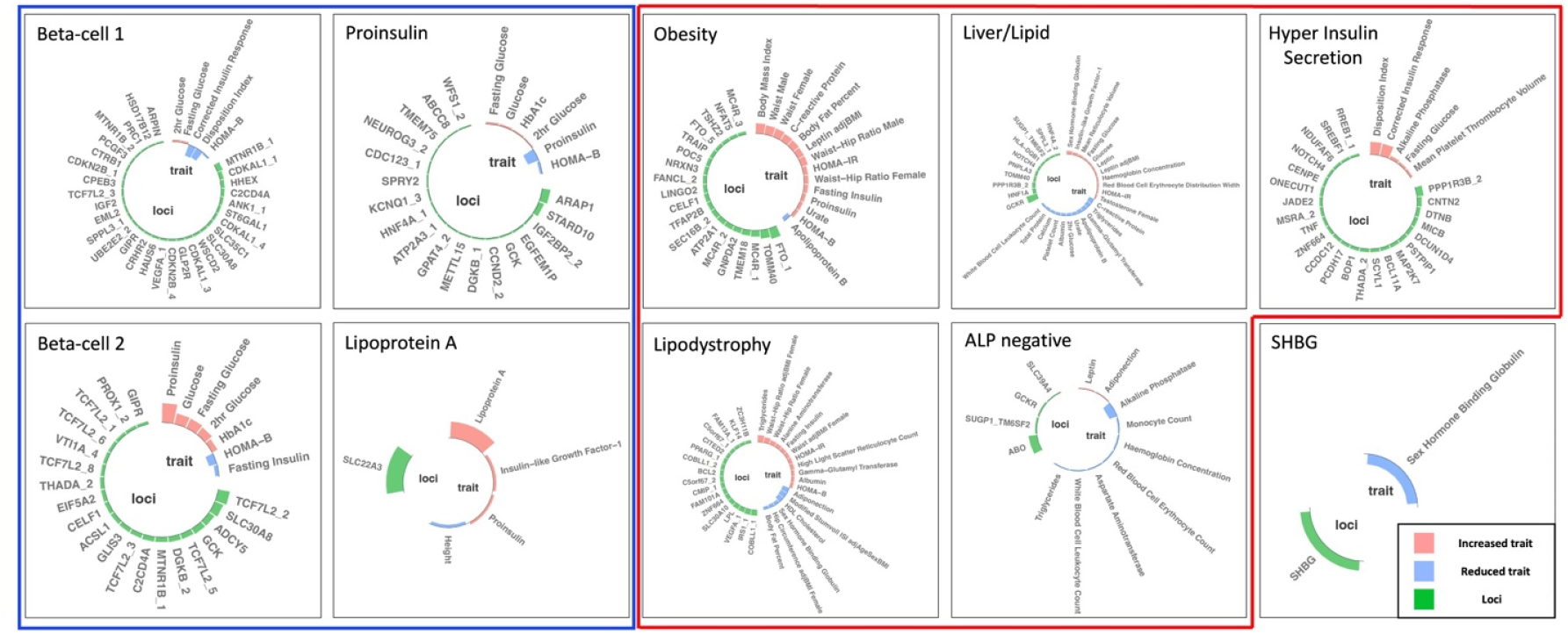
Clusters of Type 2 Diabetes Loci. Top-weighted loci and traits in each of the ten clusters are represented in circular plots. Green bars represent top-weighted loci, red bars represent increased traits, and blue bars represent reduced traits in each cluster. A maximum of 35 elements (loci and traits) based on highest weights are displayed in each cluster. Blue outline indicates clusters associated with decreased fasting insulin levels, and red outline indicates clusters associated with increased fasting insulin levels.

Of the four clusters related to insulin deficiency (Beta-cell 1, Beta-cell 2, Proinsulin, Lipoprotein A), Beta-cell 1 and Beta-cell 2 appeared to be a combination of the single Beta-cell cluster in our previous work [6], both including several well-known loci (**Table 1**). In Beta-cell 1, the top-weighted traits were decreased corrected insulin response (CIR) and decreased disposition index (DI), both indicators of reduced pancreatic beta-cell function; the most strongly weighted loci included known beta cell loci *MTNR1B, CDKAL1, HHEX, C2CD4A*, and *SLC30A8* [38–40] (**Table 1, Table S5**). Beta-cell 2 cluster was notable for having increased fasting proinsulin adjusted for fasting insulin, reduced HOMA-B and fasting insulin among the most strongly weighted traits; *TCF7L2, ADCY5, GCK, DGKB/TMEM195*, and *GLIS3* were among the top-weighted loci [41–43] (**Table S5, S6**).

The Beta-cell 1 and Beta-cell 2 clusters differed from each other with regard to the magnitude of glycemic trait effects, with Beta-cell 1 (N loci=63) having more marked association with reduced DI compared to Beta-cell 2 (beta=-0.05, *P*=3.69×10^−61^ vs beta=-0.03, *P*=9.02×10^−9^), while Beta-cell 2 (N loci=28) had a more marked association with increased fasting proinsulin adjusted for fasting insulin (beta=0.02, *P*=9.81×10^−43^ vs beta=0.006, *P*=9.81×10^−7^) and fasting glucose (beta=0.02, *P*=1.87×10^−88^ vs beta=0.008, *P*=8.78×10^−45^), compared to Beta-cell 1 cluster (**Figure 1, Table S7**). Proinsulin is a prohormone precursor to insulin, and elevated fasting proinsulin levels relative to fasting insulin levels indicates defective proinsulin processing, particularly related to beta cell stress [44, 45]. The stronger association with increased proinsulin levels for Beta-cell 2 vs Beta-cell 1 could therefore indicate that Beta-cell 2 relates more specifically to beta cell stress.

The Proinsulin cluster, also identified in our previous work, had top-weighted traits of decreased fasting proinsulin adjusted for fasting insulin and reduced HOMA-B (**Table S5, S6**). The top-weighted loci included distinct signals in the *ARAP1/STARD10* locus; beta cell-selective deletion of *StarD10* in mice has previously been shown to cause impaired insulin secretion [46]. In contrast to the other insulin deficiency clusters, the Proinsulin cluster (N loci=18) was significantly associated with decreased fasting proinsulin adjusted for fasting insulin (GWAS pPS *P*=3.51×10^−36^) (**Figure 1, Table S7**), potentially indicating a mechanism of lack of proinsulin substrate for insulin synthesis and secretion.

The Lipoprotein A cluster was novel to the present analysis and had a single highly weighted trait, increased serum lipoprotein A (Lp(a)), and a single highly weighted locus, *SLC22A3/LPA* (**Table S5, S6**). *SLC22A3/LPA* has been previously associated with serum Lp(a) levels [47] and contains the gene *LPA* encoding Lp(a). The T2D-risk-increasing allele of Lipoprotein A cluster variant (rs487152) was strongly associated with increased Lp(a) levels in the UK Biobank (GWAS pPS *P*=4.06×10^−1586^) (**Table S7**), but the underlying mechanism relating to insulin deficiency is unknown.

Of the five clusters related to mechanisms of insulin response (Obesity, Lipodystrophy, Liver/Lipid, Hyper Insulin Secretion, ALP negative), three (Obesity, Lipodystrophy, and Liver/Lipid) were also identified in our previous work, but gained additional loci (and traits) in this expanded analysis.

The Obesity cluster had most-strongly weighted traits of increased body mass index (BMI), waist circumference, percent body fat, and C-reactive protein (CRP), and key genetic signals included the well-known obesity loci *FTO* and *MC4R* [48] (**Table S5, S6**). GWAS pPS for the Obesity cluster (N loci=35) identified significant associations with increased fasting insulin (*P*=7.92×10^−22^), HOMA-IR (*P*=7.58×10^−19^), BMI (*P*=1.87×10^−1398^), percent body fat (*P*=6.94×10^−83^) and CRP (*P*=6.47×10^−260^), supporting a mechanism of obesity-mediated insulin resistance.

The Lipodystrophy cluster had top-weighted traits and loci suggestive of “lipodystrophy-like” or fat distribution-mediated insulin resistance as in our prior work [6]; these included decreased adiponectin, HDL cholesterol, and modified Stumvoll insulin sensitivity index (adjusted for age, sex and BMI), and increased triglycerides and waist-hip ratio, as well as top-weighted loci *IRS1, KLF14*, and *PPARG* [49, 50] (**Table S5, S6**). GWAS pPS for the Lipodystrophy cluster (N loci=54) was associated with increased fasting insulin (*P*=3.16×10^−43^), HOMA-IR (*P*=7.47×10^−29^) and triglycerides (*P*=1.18×10^−612^), decreased insulin sensitivity index (*P*=1.84×10^−38^) and HDL (*P*=5.19×10^−535^).

The Liver/Lipid cluster was defined by decreased triglycerides and gamma-glutamyl transferase levels, and multiple loci previously connected to hepatic lipid or glycogen metabolism, including *GCKR, HNF1A, PPP1R3B, TOMM40/APOE*, and *PNPLA3* (**Table S5, S6**) [51–56]. GWAS pPS for this cluster (N loci=11) were associated with reduced triglycerides (*P*=3.64×10^−181^) and interestingly also reduced CRP (*P*=7.75×10^−106^) and white blood cell count (*P*=1.42×10^−49^).

The two remaining insulin response clusters were novel (labeled ALP negative and Hyper Insulin Secretion). The ALP negative cluster had decreased alkaline phosphatase (ALP) level as its top-weighed trait, and the *ABO* locus as the top-weighted locus (**Table S5, S6**). GWAS pPS in this cluster (N loci=4) was associated with decreased serum ALP (*P*=1.97×10^−1431^) and triglycerides (*P*=4.49×10^−247^). The Hyper Insulin Secretion cluster included top-weighted traits of increased DI and CIR, and loci *PPP1R3B, CNTN2, DTNB, SREBF1*, and *TNF*. The Hyper Insulin Secretion GWAS pPS (N loci=32) was associated with increased CIR (*P*=1.16×10^−14^), DI (*P*=2.89×10^−14^), BMI (*P*=1.01×10^−26^), and reduced HDL (*P*=1.09×10^−110^) and SHBG (*P*=1.07×10^−100^).

The final cluster, labeled SHBG, was novel to the current work and not significantly associated with fasting insulin (GWAS pPS *P*=0.36). The cluster was driven by a single trait and locus: decreased SHBG levels and the *SHBG* locus (**Table S5, S6**). GWAS pPS in the SHBG cluster (N loci=1) was significantly associated with reduced SHBG (*P*=1.2×10^−1784^) and reduced IGF-1 (*P*=4.12×10^−13^).

### T2D Clusters differ in tissue enrichment including single cell islets

To acquire further evidence for the suspected mechanistic pathways represented by clusters and assess the biological difference between the clusters, we analyzed the top weighted loci in each cluster for enrichment of epigenomic annotations across 28 tissues. The T2D clusters displayed clearly different patterns of tissue enhancer/promoter enrichment (**Figure 3A, Table S8a**). In line with expected mechanisms, the Beta-cell 1, Beta-cell 2, and Proinsulin clusters were significantly enriched for pancreatic islet tissue (FDR<0.05). The Liver/Lipid and ALP negative clusters were significantly enriched in liver tissue (FDR<0.01). The Lipodystrophy cluster was strongly enriched for adipose tissue (FDR<0.01). Additionally, both Beta-cell 1 and 2 clusters had enrichment in adipose and the brain hippocampus (FDR<0.01). The Obesity cluster was most transcriptionally enriched in human epidermal keratinocytes (NHEK) and hASC-t3 pre-adipose cells, both at nominal significance (*P*<0.05, FDR=0.11); of note, a stigmata of insulin resistance commonly seen in obese patients with T2D is acanthosis nigricans, which is hyperpigmentation of skin driven by proliferation of epidermal keratinocytes [57].

**Figure 3.**
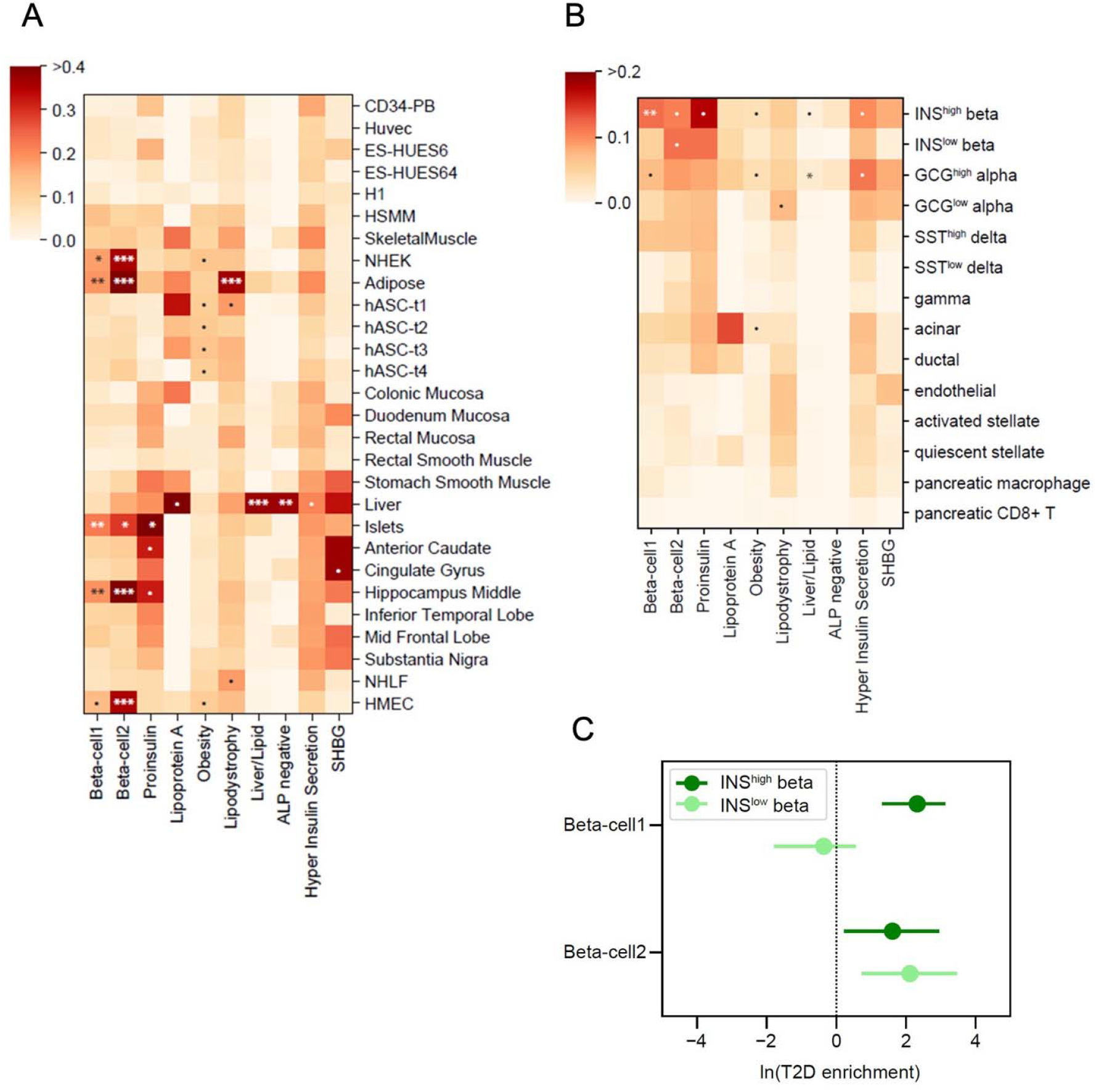
Enrichment for tissue-specific enhancers in T2D clusters. (A) Heatmap of tissue enhancer/promoter enrichment analysis result. (B) Heatmap of pancreatic islet cell enrichment analysis result. Significance was indicated as follows: *** FDR < 0.001, ** FDR < 0.01, * FDR < 0.1, • P < 0.05. (C) Comparison of Beta-cell 1 and Beta-cell 2 clusters in fgwas enrichment analysis in functional and stressed beta-cell states shown in a forest plot.

We also interrogated newly available chromatin profiles from 14.3k pancreatic islet cells, which Chiou *et al*. subsetted based on their chromatin profiles [30]. In prior work, the islets were found to have two epigenomic subsets, labeled Beta INS^high^ and Beta INS^low^, indicating high or low insulin gene (*INS)* promoter accessibility; the Beta INS^high^ islet cells were noted to be enriched for promoter accessibility for genes involved in insulin secretion, whereas the Beta INS^low^ was enriched for genes involved in stress-induced signaling response [30]. When assessing enrichment of our genetic clusters, we found that our Beta-cell 1 genetic cluster was enriched only in Beta INS^high^ cells (P=0.0001, FDR=0.0014), whereas our Beta-cell 2 genetic cluster was nominally enriched in both Beta INS^high^ and Beta INS^low^ cells (P=0.025, P=0.013, respectively, FDR=0.18 for both), (**Figure 3B, Table S8b**). Further supporting the delineation of the Beta cell loci into two separate sub-pathways, the same trend was observed in our fgwas enrichment analysis: Beta-cell 1 was significantly enriched only in INS^high^ (ln(enrichment) (95% CI) INS^high^ 2.32 (1.31-3.12); INS^low^ -0.36 (−1.79-0.55)) whereas Beta-cell 2 was significantly enriched in both single cell subsets (ln(enrichment) (95% CI), INS^high^ 1.61 (0.22 - 2.96); INS^low^ 2.11 (0.73 - 3.46)) (**Figure 3C**). Together these results supported that Beta-cell 1 and Beta-cell 2 clusters relate to distinct physiological mechanisms, with Beta-cell 2 again connected to a stress-induced pancreatic state.

Also of interest, within the pancreas single cell data, the Liver/Lipid cluster was most enriched for alpha cells, (P=0.007, FDR=0.099); alpha cells secrete glucagon, which acts to release glucose from the glycogen stores in the liver, providing further connection between these T2D genetic loci with liver function.

### T2D clusters are differentially associated with clinical traits and outcomes

To assess translation of the clusters to individuals, we generated cluster pPSs in the hospital-based MGB Biobank (N=25,419). We first confirmed that cluster pPSs were associated with expected traits in this study population both in all individuals and in just those with T2D (**Table S11**).

We next tested whether the cluster pPS were associated cardiometabolic clinical outcomes related to T2D using GWAS summary statistics: CAD, CKD, eGFR, hypertension, ischemic stroke, diabetic retinopathy, and diabetic neuropathy (**Table S4, Table S10, Figure 4a, Figure S4a**). All ten T2D clusters were associated with at least one outcome. The GWAS pPS results for eGFR highlighted the utility of cluster-specific scores, with individual clusters having more significant associations than the full set of combined T2D SNPs: increased pPSs for the Liver/Lipid, ALP negative, and SHBG clusters were associated with reduced eGFR (*P*<5×10^−4^), whereas all cluster T2D SNPs together did not reach Bonferroni-corrected statistical significance (*P*=0.03, **Table S10**). The most significant of these GWAS pPS were replicated using individual-data from MGB Biobank: increased Obesity cluster pPS had risk of hypertension, increased Lipodystrophy cluster pPS and risk of CAD, and increased Liver/Lipid cluster pPS and reduced eGFR, in all individuals with and without adjustment for T2D status (**Figure 4, Table S11**).

**Figure 4.**
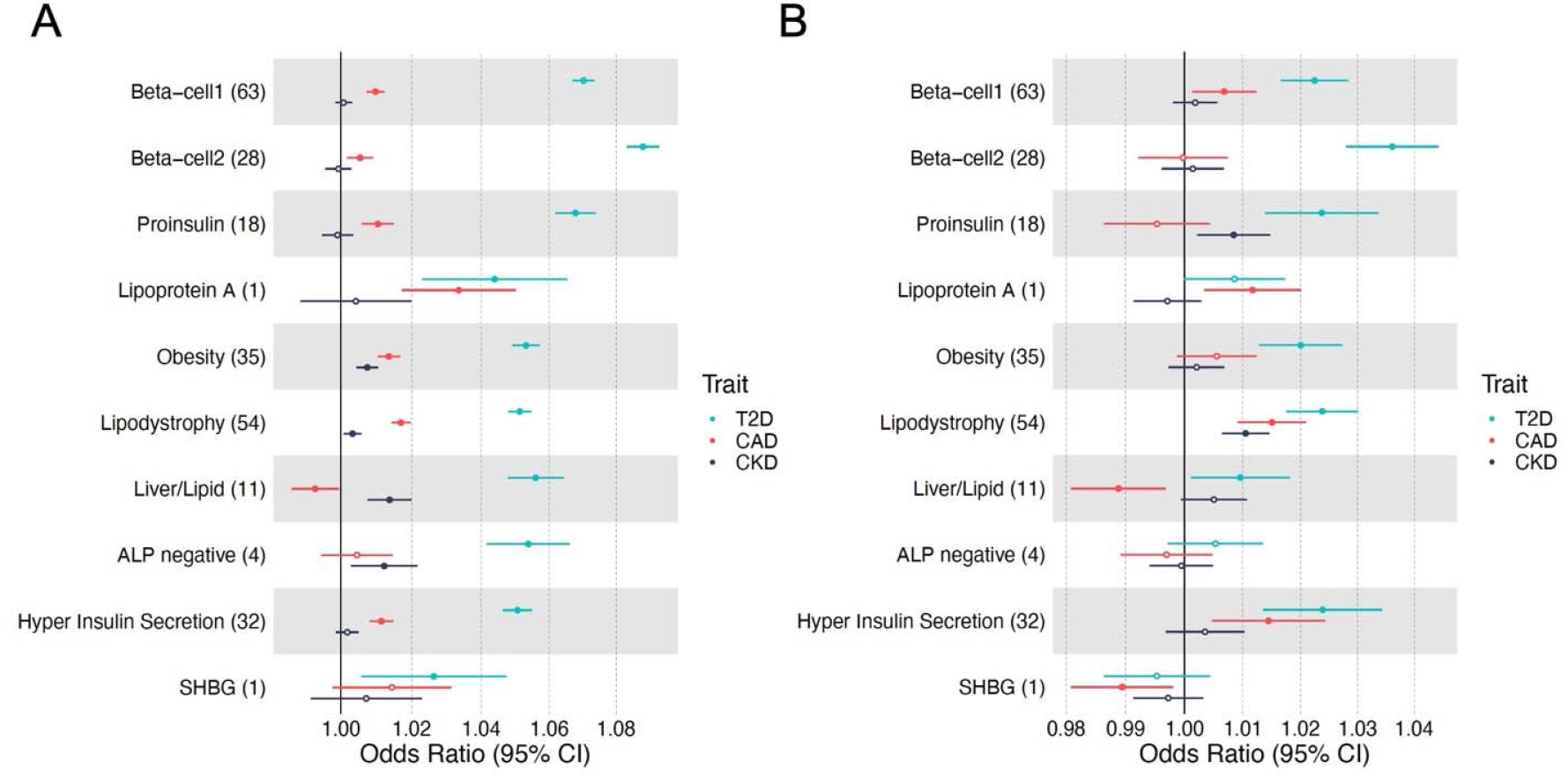
Forest plot of cluster associations with outcomes using (A) GWAS and (B) individual-level data from MGB Biobank. (A) Standardized effect sizes with 95% confidence intervals of cluster pPS-outcome associations derived from GWAS summary statistics shown in forest plot. Three metabolic outcomes (T2D, CAD and CKD, all T2D unadjusted) are displayed. The numbers in the parenthesis next to cluster names indicate the number of variants included in the analysis in each cluster. Filled points indicate *P*-values less than 0.05. (B) Associations of pPSs in individuals in the MGB Biobank with clinical outcomes are shown in forest plot. Three outcomes including T2D are displayed.

### Clusters from CAD and CKD share mechanistic pathways with T2D

To identify shared pathways among cardiometabolic outcomes and demonstrate portability of our high-throughput pipeline, we applied the same clustering approach to 219 CAD loci and 70 CKD loci. Five clusters were identified in clustering analysis of 219 CAD loci (ALP negative, Lipoprotein A, HDL negative, Cholesterol and Blood markers increased) and five clusters were identified for the 70 CKD loci (Blood markers increased, Urea increased, Reduced hematopoiesis, Beta-cell opposite and Lipoprotein A) (**Tables S12-15, Figure S5**). Based on inspection of constituent variants and traits in the clusters of T2D, CAD, and CKD, one cluster, Lipoprotein A, was shared by all three diseases. Similarly, the ALP negative cluster was shared between T2D and CAD, and the Blood markers increased cluster between CAD and CKD.

## Discussion

Novel approaches are needed to connect the currently identified hundreds of T2D genetic loci to disease pathophysiology and also accommodate the rapid pace of new loci discovery. Here, we describe expanded clustering of T2D variants, using a high-throughput pipeline for extracting and preprocessing variants from multiple GWAS datasets and generating a variant-trait association matrix. The resulting matrix consisted of 324 T2D genetic variants and 64 diabetes-related metabolic traits from publicly available GWAS datasets. By applying bNMF soft clustering to this matrix, we identified ten robust clusters of T2D variants, representing biologically meaningful mechanistic pathways.

Among the ten clusters, we replicated the five identified in our previous work of 94 T2D variants (Beta-cell, Proinsulin, Obesity, Lipodystrophy, Liver/Lipid) [6], with the Beta-cell cluster now subdivided into two distinct clusters, and also identified four additional novel clusters related to pronounced insulin secretion, levels of alkaline phosphatase, lipoprotein-A, and sex hormone-binding globulin. In contrast to our prior work, which involved manual curation of loci published in GWAS manuscripts to generate the input list of variants, the current approach allowed for use of uncurated GWAS summary statistics files and included additional newly available datasets, more than tripling the number of input genetic loci. Thus, replication of the previously identified five clusters provides strong validation of this high-throughput approach, with the newly identified clusters driven by traits or loci not available in the prior analysis.

Three of the ten T2D clusters identified in this work (Beta-cell 1, Beta-cell 2, and Proinsulin) clearly related to pancreatic beta-cell function, with the two Beta-cell clusters differing from the Proinsulin cluster with regard to the direction of association with fasting proinsulin adjusted for fasting insulin. All three clusters were enriched in pancreatic islet tissue enhancers and promoters in the epigenomics analysis. Additionally, loci in the Beta-cell 1 cluster were significantly enriched for a subset of single beta cells predicted independent of our work based on RNA transcript levels to represent a normal state of pancreatic beta cell function, whereas loci in Beta-cell 2 cluster had a unique signal of enrichment for single beta cells predicted to be in a stressed state [30]; these functional distinctions between Beta-cell 1 and 2 supported our independent approach of phenotypically informed clustering T2D loci.

Three other T2D genetic clusters (Obesity, Lipodystrophy, Liver/Lipid) replicated findings from our prior work related to pathways of insulin resistance, gaining additional loci and traits compared to the prior analysis. Loci in these three clusters were most enriched for enhancers in tissues for the suspected mechanisms: pre-adipocytes, adipocytes, and liver tissue, respectively. The distinction between fat accumulation in the Obesity cluster and abnormal fat compartmentalization in the Lipodystrophy cluster may be supported by the differential enhancer enrichment shown for different developmental stages of the same adipocyte lineage.

In addition to a second Beta-cell cluster, there were four newly identified T2D genetic clusters from this work: ALP negative (containing the *ABO* locus), Lipoprotein A, SHBG, and Hyper Insulin Secretion.

The ALP negative cluster seen for T2D and CAD was driven by reduced serum alkaline phosphatase levels and the *ABO* locus. Isoforms of alkaline phosphatase have been shown to vary in level by blood group [58]. The *ABO* locus and blood type have previously been connected to T2D [59–61] and CAD [62] risk, but the causal mechanisms are not fully understood.

The Lipoprotein A cluster seen for T2D, CAD, and CKD all included the locus (*SLC22A3*/*LPA* tagged by rs487152) and biomarker Lp(a), pointing to a genetic pathway leading to increased Lp(a) levels and also increased risk of T2D, CAD, and CKD. The relationship between Lp(a) and cardiometabolic disease is complex, and genetic interrogation of *LPA* has been complicated by the fact that plasma concentration of Lp(a) is influenced by kringle IV type 2 (KIV-2) repeats in addition to other genetic variation [63]. While epidemiological studies have connected elevated Lp(a) levels with increased risk of CAD and CKD [64, 65], an inverse association has been reported for T2D [66, 67]. Our genetic findings for T2D therefore indicate that there are likely multiple pathways impacting Lp(a) level that may have differential effects on T2D risk.

The SHBG cluster was also driven by a single locus and biomarker. Our results point to a genetic pathway whereby alteration of the *SHBG* locus leads to reduced SHBG levels and increased T2D risk, which was consistent with previous epidemiological and genetic studies indicating that low circulating levels of SHBG were causally related to increased risk of T2D in women and men [68, 69].

We assessed the impact of cluster pPSs in individuals, finding that individuals with increased cluster pPS had significant associations with clinical traits and outcomes (**Figure 4, Table S9, S11**), supporting prior findings for the original five clusters [70]. These results point to marked heterogeneity in T2D genetic associations that could be missed without delineation of loci into clusters and also suggest important physiological differences between clusters. At the same time, the effect sizes of the present pPSs on clinical outcomes were likely too small to be of clinical utility at the individual-level.

The strengths of this study include the high-throughput approach for preprocessing variants and traits from multiple GWAS datasets in a semi-automated way. This method can also be readily applied to other diseases beyond T2D to identify key pathways and code has been made freely available. We included here application of the pipeline to CAD and CKD, demonstrating transferability of the approach and potential shared pathways among the three cardiometabolic outcomes. Limitations include clustering of only available phenotypes from GWAS. It is possible that additional pathways exist that are not captured using the set of traits included in the analysis. Additionally, due to methodological limitations and data availability we have focused on GWAS datasets from populations of European ancestry, although we are actively pursuing application of this method in non-European populations through additional efforts. It is worth noting that bNMF generates weights for all included elements in the matrix, and it is not known how best determine a cut-off threshold for cluster membership; we have applied a reasonable strategy to maximize signal to noise. Finally, whether the associations of specific genetically derived clusters with metabolic traits and outcomes remain constant throughout the disease course has not been examined in this cross-sectional analysis.

In summary, we have identified ten robust genetic clusters pointing to mechanistic pathways of T2D using a high-throughput clustering pipeline of GWAS summary statistics. These clusters displayed tissue-specific enrichment patterns even within single cell pancreatic tissue and could be used to generate pPSs that stratify patients genetically with distinct associations with clinical features and cardiometabolic outcomes. We demonstrate that our approach can be applied to other complex diseases, with identification of shared genetic pathways between T2D, CAD, and CKD. Thus, we contribute to further delineation of cardiometabolic disease genetic pathways using a data-driven approach informed by physiology.

## Data Availability

All data produced in the present study are available upon reasonable request to the authors

## Description of Supplemental Data

Supplemental Data include five figures and fifteen tables.

## Data and Code Availability

Code for variant pre-processing, bNMF clustering, and basic visualizations is available at https://github.com/gwas-partitioning/bnmf-clustering.

## Contribution Statement

MSU, JCF, MG, and JC conceived the research question. MSU, JK, MG, KS, KEW, JC, and KG conceived the methodology, which included implementation of the clustering computational pipeline. HK, JC, MG, TM, JMM and MSU curated the data. HK, KS and JC conducted the analysis and visualized the results. HK, JC, and MSU wrote the initial draft of the paper and incorporated co-author comments. KEW, KS, JBC, TM, MG, JMM, SK, JCF, KG and AKM provided feedback on the analysis, and critically reviewed the manuscript. All co-authors approved the final version of the paper. MSU is the guarantor of this work and, as such, had full access to all the data in the study and take responsibility for the integrity of the data and the accuracy of the data analysis.

## Web Resources

Interactive results are viewable on the Common Metabolic Disease Knowledge Portal (https://hugeamp.org/).

