## Supplemental Figures for "High-throughput Genetic Clustering of Type 2 Diabetes Loci Reveals Heterogeneous Mechanistic Pathways of Metabolic Disease"

Supplementary Figures

**Supplementary Figure 1. Overview of variant preprocessing pipeline**

(A) Flowchart of the steps implemented in the variant preprocessing pipeline. Steps include: 1) extract variants from multiple GWAS datasets of an outcome of interest, 2) remove variants that do not pass a Bonferroni p-value from the largest GWAS dataset of the outcome, 3) find proxy variants for variants that are multi-allelic, ambiguous, or have low trait counts, 4) LD-prune to ensure independent signals and 5) align variants to risk increasing alleles of the outcome of interest. (B) Flowchart of trait preprocessing. Steps include: 1) filter trait GWAS by minimum sample size, 2) filter trait GWAS by a minimum Bonferroni p-value across the selected variants, 3) filter by correlation between traits and 4) generate a variant by trait association matrix.

| **a**  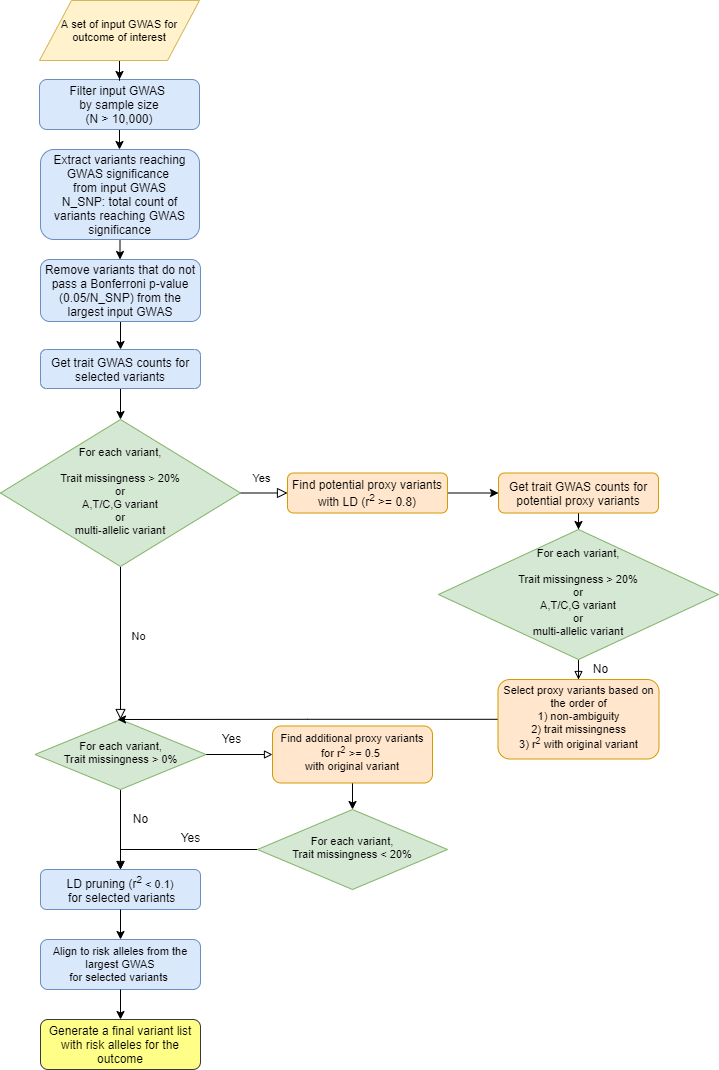 |
| --- |

| **b**  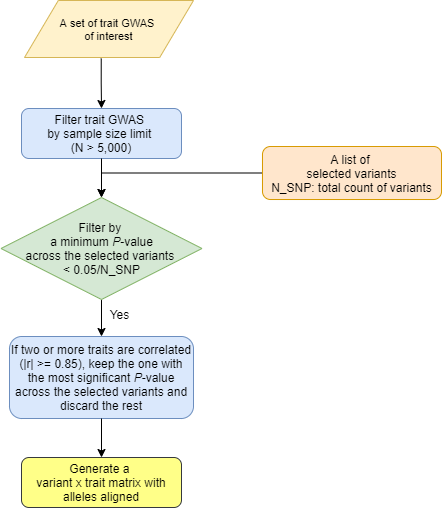 |
| --- |

**Supplementary Figure 2. Method for determining cluster weight cutoff**

The method of determining cluster weight cutoff shown in plot. First, the weights were aggregated from all the clusters and plotted in a descending order (x axis: scaled count, y axis: scaled cluster weight). Then a line (blue) was fitted to the top 1% of the weights and another line (red) to the bottom 80% of the weights. The point where the distance to the first line (blue) becomes shorter than the second line (red) is determined to be the cutoff for cluster weights.

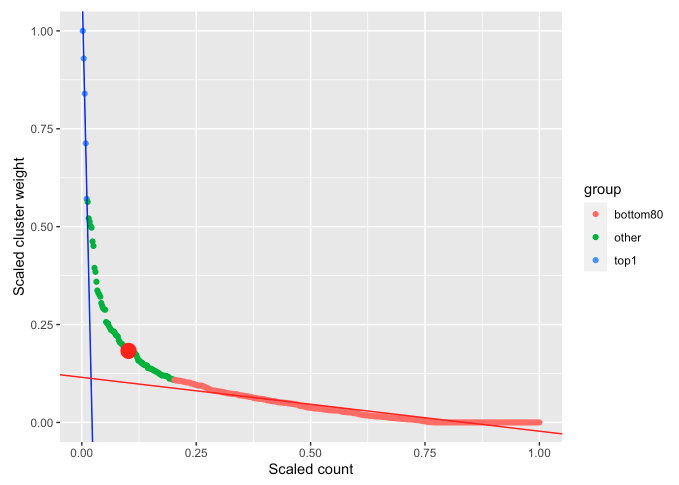

**Supplementary Figure 3. Cluster-defining characteristics**

Standardized effect sizes of cluster pPS-trait association derived from GWAS summary statistics shown in bar plots. The black vertical line (y-axis) shows an estimate of 0, representing no association between the cluster pPS and trait. A subset of discriminatory traits are displayed. Bars pointing towards the right side represent positive cluster-trait associations, whereas those pointing towards the left side represent negative cluster-trait associations. Asterisks represent significant cluster-trait associations below a Bonferroni P-value.

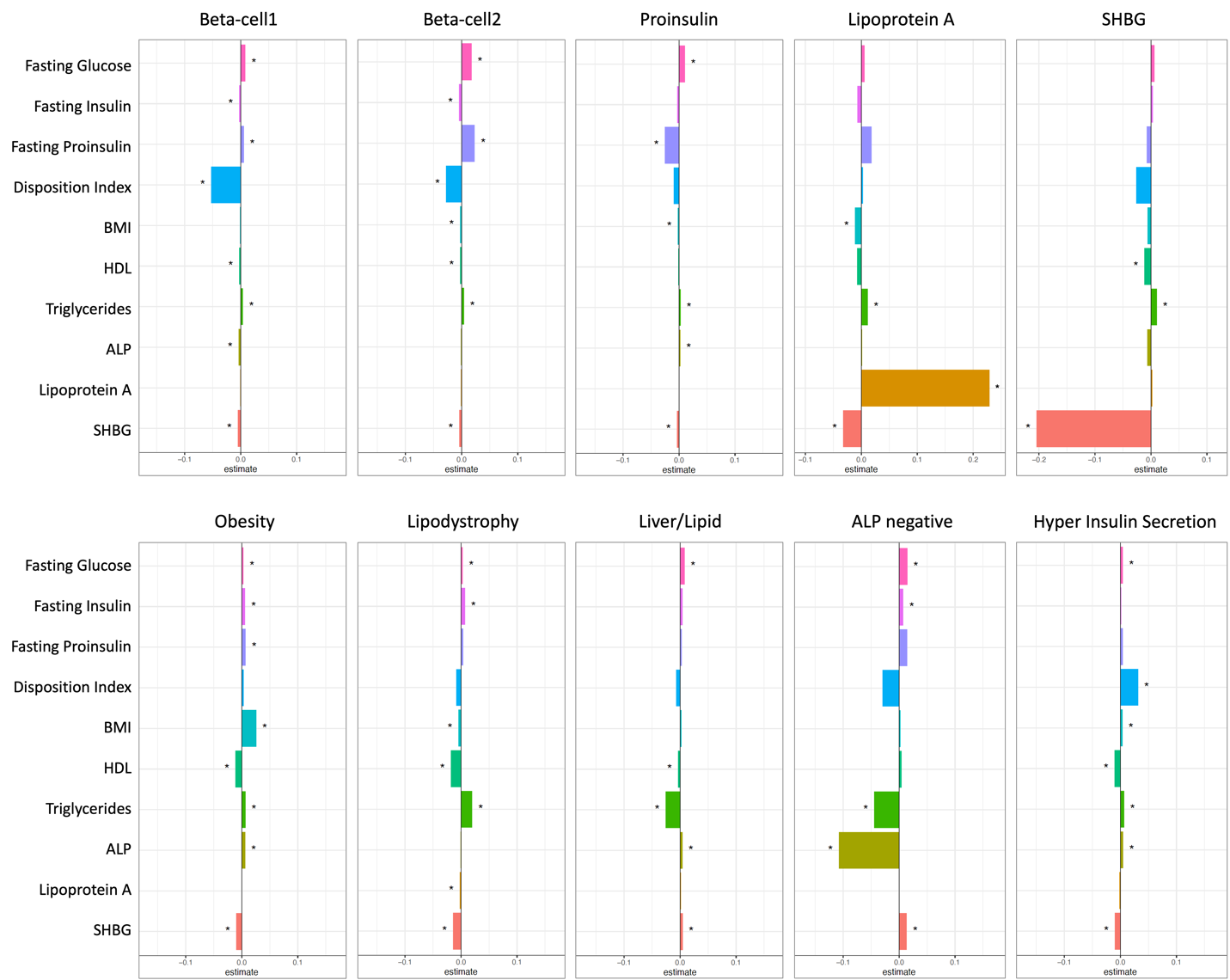

**Supplementary Figure 4. Forest plots of associations with clinical traits using a) GWAS and b) Individual-level data from MGB Biobank**

**(a)** Standardized effect sizes with 95% confidence intervals of cluster pPS-trait associations derived from GWAS summary statistics shown in forest plot. Three glycemic traits (triglycerides, HbA1c, BMI) are displayed. “All SNPs” include all the variants that are top-weighted in at least one cluster. **(b)** Associations of pPSs in individuals adjusted for T2D status in the MGB Biobank with glycemic traits included in the clustering analysis are shown in forest plot. “GRS total” is the sum of pPS using betas from T2D GWAS (DIAMANTE) of all top-weighted variants in ten clusters. The numbers in the parenthesis next to cluster names indicate the number of variants included in the analysis in each cluster. Hollow points indicate the *P*-value is equal to or greater than 0.05, and otherwise filled.

| **a**  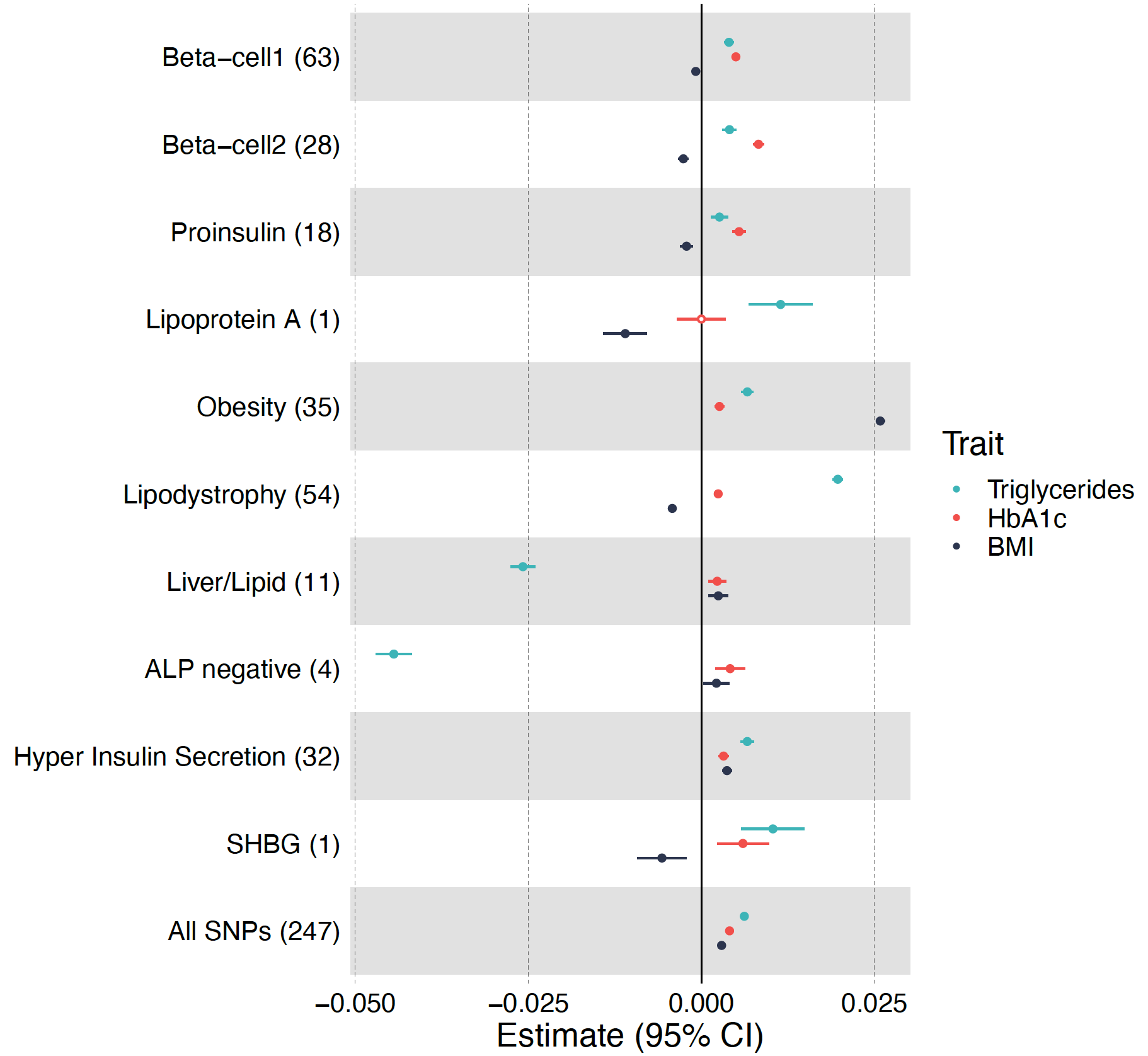 |
| --- |

**b**

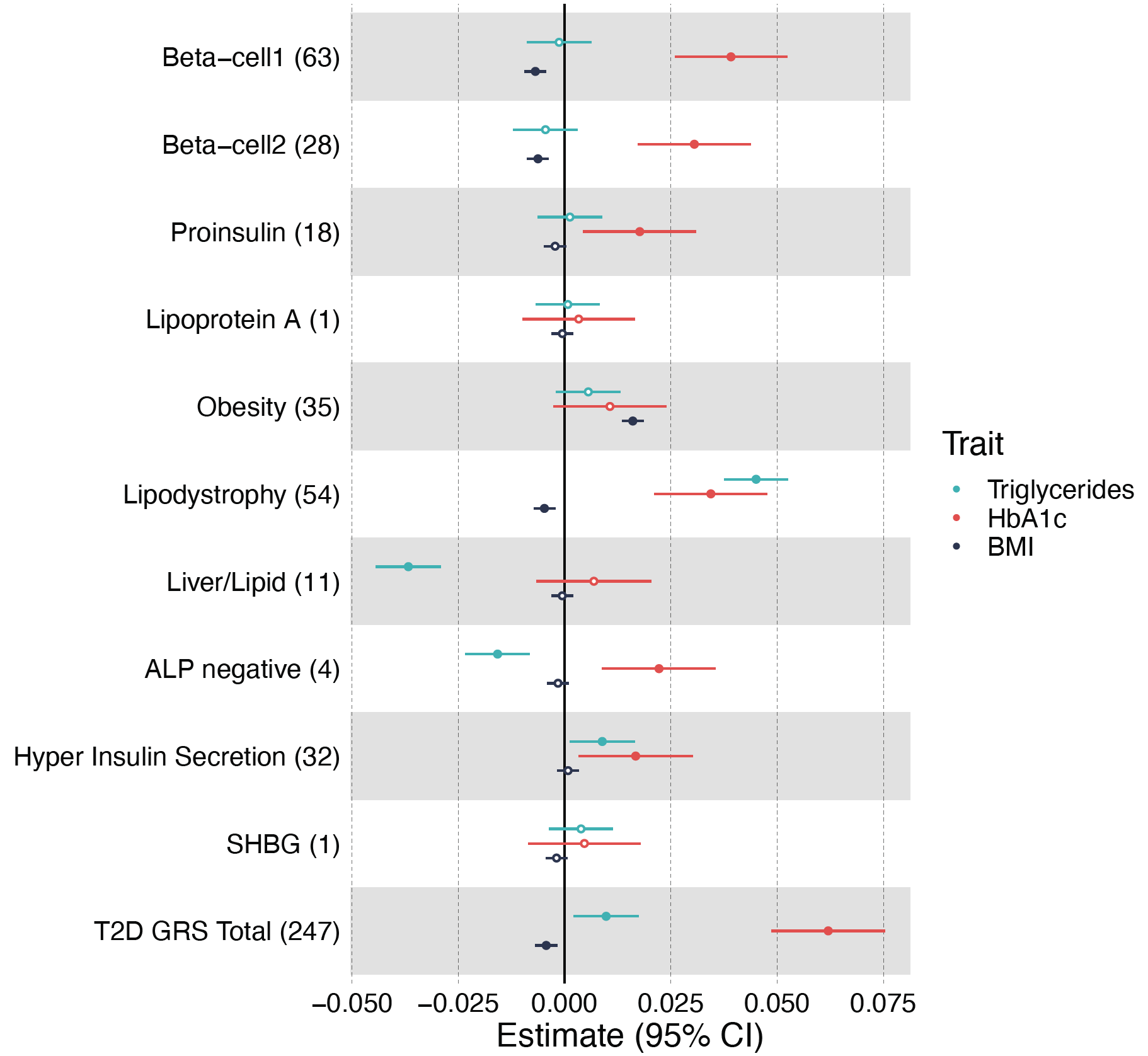

**Supplementary Figure 5. Clusters of a) CAD loci and b) CKD loci**

**(a)** Top-weighted loci and traits in each of the five CAD clusters are represented in circular plots. Green bars represent top-weighted loci, red bars represent increased traits and blue bars represent reduced traits in each cluster. **(b)** Top-weighted loci and traits in each of the five CKD clusters are represented in circular plots.

| **a**  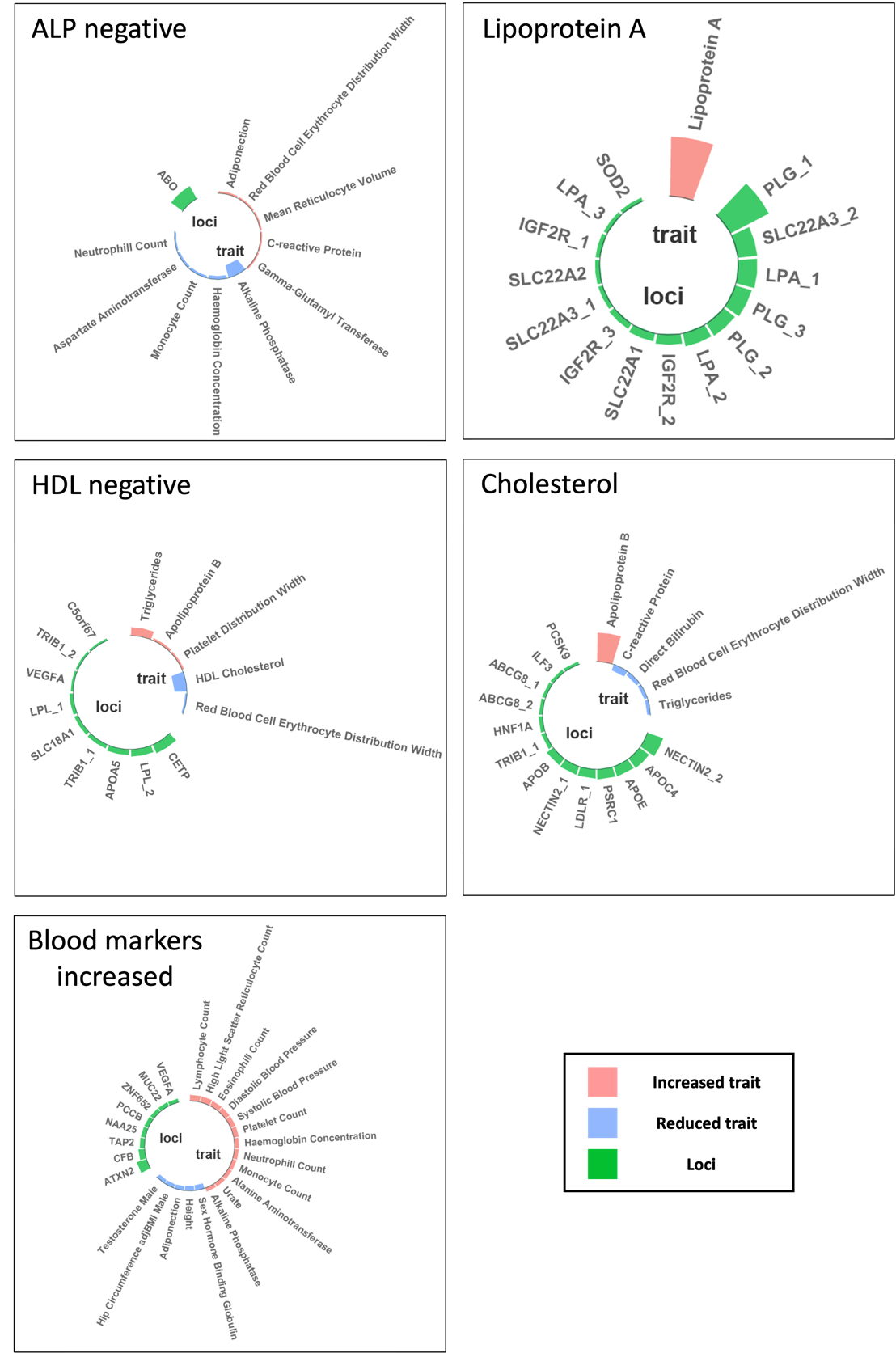 |
| --- |

| **b**  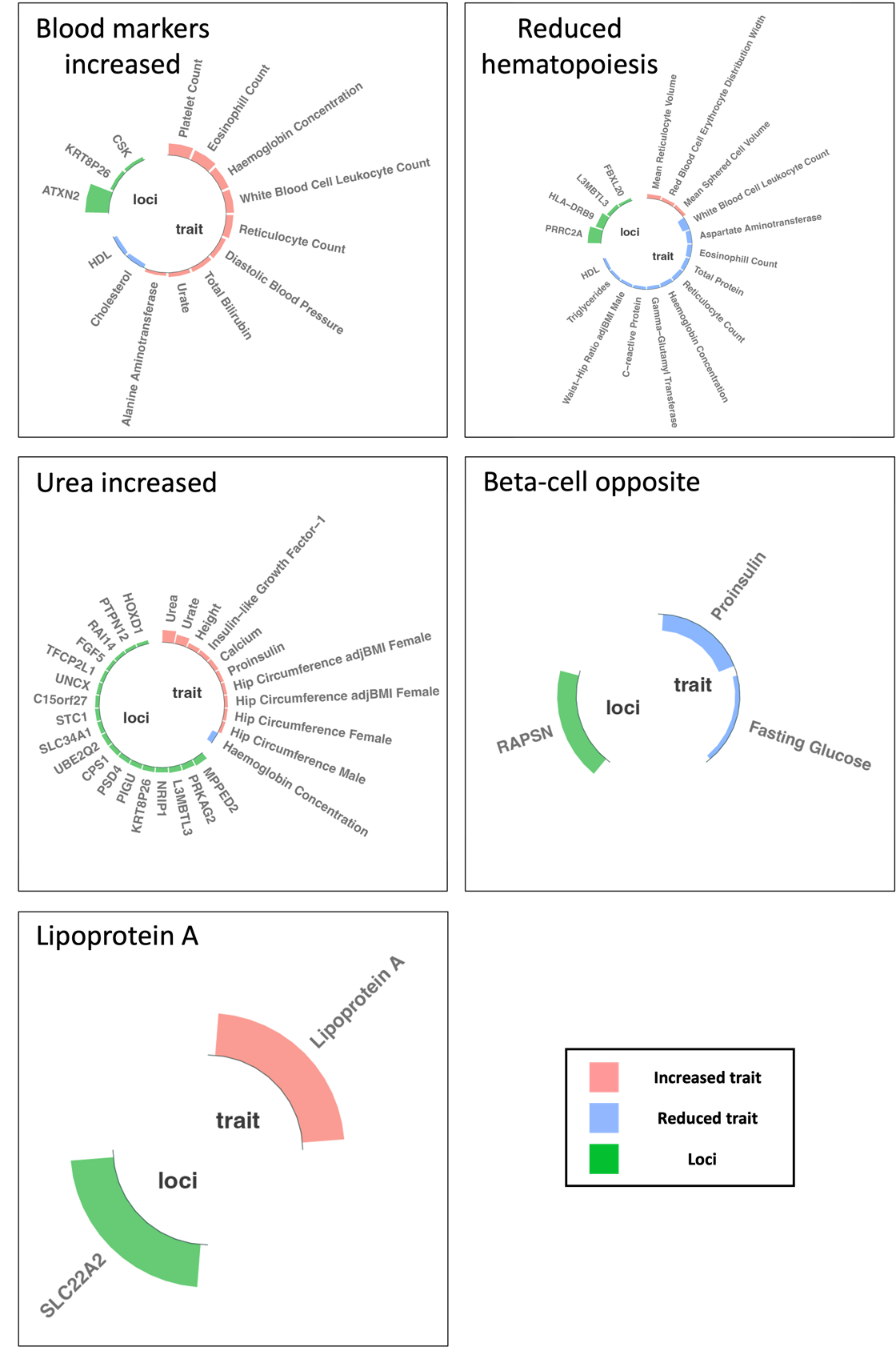 |
| --- |
