## Supplemental Methods for "High-throughput Genetic Clustering of Type 2 Diabetes Loci Reveals Heterogeneous Mechanistic Pathways of Metabolic Disease"

**Supplementary Methods**

*Variant selection*

To obtain a comprehensive set of independent genetic variants associated with T2D, we gathered T2D GWAS, exome-chip, and whole-genome sequencing summary statistic datasets deposited in the AMP-Common Metabolic Disease Knowledge Portal (CMDKP) [1]. We required the GWAS to have sample sizes larger than 10,000 to reduce false positive associations and focused on studies of predominantly European ancestry to minimize heterogeneity across studies and reduce artifactual clustering results. (Inclusion of GWAS from more than one ancestral population may lead to either genetic variants not being present due allele frequency differences across populations and/or lead to the results from the clustering method artifactually driven by patterns reflective of ancestry rather than disease biology.) Thirteen GWAS datasets were included as input for identifying T2D genetic loci for clustering (**Table S1**).

From the selected thirteen T2D GWAS datasets, we first extracted 21,666 variants reaching genome-wide significance (*P* < 5×10^−8^) and then ensured variant signals replicated in the largest of these studies at a Bonferroni significance level (*P*-value<0.05/21,666 tests). Next we identified the set of T2D variants (N=16,074) that were multi-allelic, ambiguous (A/T or C/G), or represented in less than 80% of trait GWAS datasets, and found proxy variants in linkage disequilibrium (LD) using HaploReg v4.1 [2, 3]. One proxy variant was selected to represent each locus based on prioritization of 1) non-ambiguity, 2) trait GWAS representation and 3) strong LD (r^2^ ≥ 0.8) with the initial variant. Ambiguous variants were avoided to reduce the chance of using the incorrect allele when extracting data from multiple summary statistic files.

This expanded set of T2D-associated variants was then LD-pruned based on LD data of European ancestry (CEU) using the LDlinkR package in R by LD Link [3–5]. We performed stringent LD-pruning of variants using r^2^ < 0.1; when performing LD-pruning, variants with lower *P*-values for T2D were preferentially retained. Following LD-pruning, the final LD-pruned list of T2D variants consisted of 324 variants (**Table S2**). For each variant, we identified the T2D risk allele with odds ratio (OR) > 1 using the largest available T2D GWAS and used this allele in assessing associations with traits.

A total of six GWAS and exome-chip datasets [6–8] were used as input for identifying coronary artery disease (CAD) genetic loci. For chronic kidney disease (CKD), 39 GWAS summary statistics results were queried, which included CKD GWAS as well as diabetic kidney disease (DKD), end-stage renal disease (ESRD), estimated glomerular filtration rate (eGFR) and cystatin C [9–14]. The additional kidney GWAS were included to expand the number of loci; variants were included only if at least one of their associations with CKDGen, DNCRI, and SUMMIT CKD GWAS [11–13] was significant at Bonferroni *P*-value cutoff of 0.05/N_initial_variants (N=2,269) in order to ensure correct risk allele alignment. The aforementioned variant selection procedure was applied to CAD and CKD variants identified from the input datasets, selecting 219 variants for CAD and 70 for CKD.

*Trait selection*

For trait selection, we utilized summary statistics available for GWAS of glycemic traits, anthropometric traits, vital signs, and additional laboratory measures in the AMP-CMDKP. We restricted the analysis to GWAS of continuous traits with sample sizes larger than 5,000 to reduce false positive associations and selected studies from predominantly European populations to minimize heterogeneity, as noted above. Additionally, to incorporate additional potentially relevant biomarkers, we included GWAS summary statistics of serum biomarkers available from the UK Biobank [10]. Our goal was to let the genetics guide which traits were included in the clustering analysis, and thus traits were used only if the minimum *P*-value across the final set of variants was lower than a Bonferroni *P*-value cutoff of 0.05/N_final_variants (N=324); therefore all the traits included in the clustering analysis had robust associations with at least one T2D variant. If two or more traits were highly correlated (|r| ≥ 0.85), we kept the one with the most significant *P*-value across the selected variants and discarded the rest. Among an initial set of 75 trait GWAS datasets, two traits were dropped during the minimum *P*-value filtering process and nine were dropped due to high correlation with other traits for the selected T2D variant set, leaving 64 traits (**Table S3**).

For the selected lists of variants and traits, we utilized the GWAS summary statistics to generate a matrix of standardized Z-scores, choosing the T2D risk-increasing allele for each variant and dividing the estimated regression coefficient beta by the standard error. To account for the differences in sample size across trait GWAS studies, we scaled the standardized Z-scores in a two-step process: each value was divided by the square root of the sample size for each variant in each trait GWAS, then all elements were multiplied by the mean of square root of median sample size across all SNPs in each GWAS.

**bNMF clustering**

The variant-trait association matrix Z (m by n, m: # of variants, n: # of traits) was constructed as above. We then generated a non-negative input matrix X (2m by n) by concatenating two separate modifications of the original Z matrix: one containing all positive standardized Z-scores (zero otherwise) and the other all negative standardized Z-scores multiplied by -1.

The bNMF procedure factorizes X into two matrices, W (2m by K) and H^T^ (n by K), as X ~ WH with an optimal rank K, corresponding to the association matrix of variants and traits to the number of clusters, respectively. While conventional nonnegative matrix factorization (NMF) requires the desired model order K as an input, bNMF determines an optimal K which best balances between an error measure ||X−WH||^2^ and a penalty for model complexity derived from a nonnegative half-normal prior for W and H [15–17]. Furthermore, bNMF iteratively regresses out irrelevant components in representing X with an automatic relevance determination technique, which enables an optimal inference for the number of clusters K. The key features for each cluster are determined by the most strongly associated traits, a natural output of the bNMF approach. bNMF algorithm was performed in R Studio for 1,000 iterations with maximum number of cluster K set to 20, and the maximum posterior solution at the most probable K was selected for downstream analyses. The output of this clustering consists of matrices of cluster-specific weights for each variant (W) and trait (H) [18].

To define a set of strongest-weighted variants in each cluster and maximize the signal to noise ratio of weights, we developed a method to determine a cluster weight cutoff for the clusters (**Figure S2**). This involved aggregating the weights from all the clusters and plotting them in descending order. We fitted a line to the top 1% of the weights and another line to the bottom 80% of the weights. We identified the point where the distance to the first line became shorter than to the second line, and made this the cutoff for cluster weights. For T2D, cluster weight cutoff was thus set at 0.832.

**Cluster associations with relevant phenotypes using GWAS summary statistics**

To better characterize each cluster, particularly with regard to the associations of the loci with glycemic traits included in the clustering process, we generated GWAS-partitioned polygenic scores (GWAS pPS) for each cluster, utilizing inverse-variance weighted fixed effects meta-analysis of GWAS summary statistics. For these analyses, the set of strongest-weighted variants above the weight cutoff for each cluster (as described above) were included in the model. We performed these meta-analyses with the dmetar package in R [19] using GWAS summary statistics.

Additionally, we applied this same approach to test cluster associations with relevant cardiometabolic outcomes studied in GWAS. As opposed to above, these outcomes were independent of the traits included in the bNMF clustering analysis. We tested associations between each cluster and seven relevant cardiometabolic disease outcomes; coronary artery disease (CAD), chronic kidney disease (CKD), estimated glomerular filtration rate (eGFR), hypertension, ischemic stroke, diabetic retinopathy, and diabetic neuropathy (**Table S4**). The significance threshold was set to 0.05/(7×K), representing a Bonferroni correction for 7 outcomes and K clusters.

**Functional annotation and enrichment analysis**

At each locus, we calculated approximate Bayes Factors (aBF) for all variants 500 kb upstream and downstream with r^2^ ≥ 0.1, with the index variant (100% credible set) from effect size estimates and standard errors, using the approach of Wakefield [20]. We then calculated a posterior probability for each variant by dividing the aBF by the sum of all aBF in the credible set.

We obtained previously published 13-state ChromHMM [21] chromatin state calls for 28 cell types, excluding cancer cell lines [22]. For each cell type, we extracted chromatin state annotations for enhancer (Active Enhancer 1, Active Enhancer 2, Weak Enhancer, Genic Enhancer) and promoter (Active Promoter) elements. We also compiled candidate cis-regulatory elements (cCREs) for 14 cell types and subtypes from published single cell chromatin accessibility datasets [23, 24].

We assessed enrichment of annotations within clusters by overlapping 100% credible set variants for signals in each cluster with cell type epigenomic annotations (chromatin states and cCREs). We calculated cell type probabilities for each cluster by summing the posterior probabilities of variants in cell type enhancers or promoters, divided by the number of signals in the cluster. We derived significance for cell type probabilities for each cluster using a permutation-based test. We permuted signals and cell type labels within each cluster and then recalculated cell type probabilities, as above. We then used cell type probabilities derived from 10,000 permutations as a background distribution and performed a one-tailed test to ascertain significance for each cell type.

We also assessed epigenomic enrichment in single cell pancreatic tissue using a second method. As previously described [25], we subset loci from the Beta-cell 1 and 2 clusters, annotated variants using cCREs from INS^high^ and INS^low^ beta cells, and applied fgwas [25] in the fine mapping mode. We considered annotations significantly enriched if the lower bound of the 95% confidence interval of the natural log enrichment was greater than 0.

**Partitioned Polygenic Score (pPS) analysis in the Mass General Brigham Biobank**

The Mass General Brigham (MGB) Biobank (formerly Partners Biobank) provides banked samples (plasma, serum, DNA and genomics data) collected from more than 120,000 consented patients seen at hospitals and clinics across the MGB system, including Brigham and Women's Hospital, Massachusetts General Hospital, Massachusetts Eye and Ear Infirmary, Faulkner Hospital, Newton-Wellesley Hospital, McLean Hospital, North Shore Medical Center and Spaulding Rehabilitation Hospital, all in the Boston area of Massachusetts [26, 27]. Patients are recruited at clinical care appointments at more than 40 sites and clinics, and also electronically through the patient portal at MGB. Biobank subjects provide consent for the use of their samples and data in clinical research. Written consent was provided by all study participants. Approval for analysis of Biobank data was obtained by the MGB IRB, study 2016P001018.

T2D status was defined based on algorithmically defined phenotypes developed by the Biobank Portal team using both structured and unstructured electronic medical record data and clinical, computational, and statistical methods [28]. Cases were selected by this curated phenotype to have T2D with PPV of 99% and required to be of at least age 35 to further minimize misclassification of T2D diagnosis. Additional phenotypic data (laboratory measures, vital signs, and anthropometric measures) were extracted, from which we generated median values over the most recent 5 years available within the years of 2015-2020.

Up to 36,000 samples were genotyped using three versions of the Biobank SNP array offered by Illumina that is designed to capture the diversity of genetic backgrounds across the globe. The first batch of data was generated on the Multi-Ethnic Genotyping Array (MEGA) array, and the second, third, and fourth batches were generated on the Expanded Multi-Ethnic Genotyping Array (MEGA Ex) array. All remaining data were generated on the Multi-Ethnic Global (MEG) BeadChip. The genotyping data were harmonized and quality controlled with a three-step protocol, including two stages of SNP removal and an intermediate stage of sample exclusion. The exclusion criteria for genetic variants were 1) missing call rate ≥ 0.05, 2) significant deviation from Hardy-Weinberg equilibrium (*P* ≤ 10^-20^ for the entire cohort), and 3) minor allele frequency (MAF) < 0.001. The exclusion criteria for samples were 1) gender discordance between the reported and genetically predicted sex, 2) subject relatedness (pairs with ≥ 0.125, from which we removed the individuals with the highest proportion of missingness), 3) missing call rates per sample ≥ 0.02, and 4) population structure showing more than four standard deviations within the distribution of the study population, according to the first four principal components (PCs). Phasing was performed with SHAPEIT [29] and then imputed with the Haplotype Consortium Reference Panel [30] using the Michigan Imputation Server [31].

We performed individual-level analyses on samples restricted to individuals from European ancestry based on self-reported ancestry and genetic PC’s, totaling 25,419 individuals. SNPs were included in genetic risk scores as allele dosages. All SNPs were genotyped or imputed with high quality (r^2^ values > 0.95). T2D partitioned polygenic scores (pPSs) for each cluster were generated by multiplying a variant's genotype dosage by its cluster weight. For each cluster pPS, only the top-weighted variants were included, as defined above. Logistic and linear regression were performed in R v3.6.2, adjusting for age, sex, and PC’s.
